# Heart Rate Modulation and Clinical Improvement in Major Depression: A Randomized Clinical Trial with Accelerated Intermittent Theta Burst Stimulation

**DOI:** 10.1101/2025.04.06.25325324

**Authors:** Jonas Wilkening, Henrike M. Jungeblut, Ivana Adamovic, Vladimir Belov, Peter Dechent, Lara Eicke, Niels Hansen, Vladislav Kozyrev, Lara E. Marten, Yonca Muschke, Caspar Riemer, Knut Schnell, Asude Tura, Melanie Wilke, Fabian Witteler, Jens Wiltfang, Anna Wunderlich, Valerie Zimmeck, Anna Zobott, Carsten Schmidt-Samoa, Tracy Erwin-Grabner, Roberto Goya-Maldonado

**Author notes:** Corresponding author: PD Dr. Roberto Goya-Maldonado Laboratory of Systems Neuroscience and Imaging in Psychiatry (SNIP-Lab, www.sniplab.uni-goettingen.de) Department of Psychiatry and Psychotherapy University Medical Center Göttingen (UMG) Von-Siebold Str. 5, 37075 Göttingen.

## Abstract

Intermittent theta burst stimulation (iTBS) is a promising non-invasive treatment for major depressive disorder (MDD), though significant variability exists in patient responses. This quadruple-blind, sham-controlled crossover randomized clinical trial investigated whether clinical improvement was influenced by the spatial selection of stimulation sites or early cardiac rhythm modulations. Seventy-five MDD patients (33 women) were randomized to either personalized stimulation sites— identified via the strongest negative functional connectivity between the left dorsolateral prefrontal cortex (DLPFC) and the default mode network (DMN)—or fixed stimulation at the F3 position of the 10-20 EEG system. Electrocardiograms throughout stimulation monitored heart rate and its variability, focusing on heartbeat deceleration (RR slope) and changes in the root mean square of successive differences between heartbeats (RMSSD). Findings demonstrated that a higher RR slope within the first 45 seconds of stimulation predicted greater clinical improvement at six weeks, while a lower RMSSD change during the first 270 seconds correlated with improvement at one week. However, no significant differences in outcomes were observed between personalized and fixed stimulation sites. This study identified a significant relationship between early heart rhythm modulations and the antidepressant effects of accelerated iTBS in MDD. Although the spatial determination of stimulation sites did not enhance clinical improvement, iTBS-induced changes in cardiac rhythm during early sessions may develop into valuable biomarkers for stratifying patients, enabling more personalized and effective treatment strategies in the future.

## Introduction

Major depressive disorder (MDD) is one of the most common psychiatric disorders and the leading cause of disability worldwide (1). With a lifetime prevalence of 16-20%, MDD treatment is particularly challenging for approximately one-third of patients who experience treatment-resistant depression (TRD). These patients fail to respond to two or more antidepressant medication trials of adequate dose and duration (2). In this context, repetitive transcranial magnetic stimulation (rTMS) has gained attention as an effective treatment modality (3, 4). More recently, advances in stimulation protocols such as intermittent theta burst stimulation (iTBS) (5), along with the expansion of neurostimulation in clinical settings, have encouraged the application of rTMS as a first-line of treatment for MDD (6, 7). Although access to and familiarity with iTBS has increased, clinical outcomes remains extremely variable and difficult to predict. In the field of precision medicine, neurobiological predictors have been proposed as potential markers of clinical improvement in patients undergoing rTMS. For example, the selection of customized stimulation sites based on individual functional connectivity and the identification of heart rhythm modulations at the onset of stimulation are very promising candidates (8–11). Thus, in this randomized clinical trial (RCT), we prospectively evaluated the clinical improvement of spatially personalized stimulation sites as well as subsequent heart rate (HR) and HR variability modulations. Both strategies suggest that the degree of communication from the site of stimulation in the left dorsolateral prefrontal cortex (DLPFC) to the subgenual anterior cingulate cortex (sgACC) is likely implicated in clinical improvement (12–14).

The most studied and successful coil position for the treatment of clinical depression has been in the left DLPFC (15). Historically, the position site was determined using the “5cm rule” or “7cm rule”, referring to the distance from the point where the resting motor threshold (RMT) was established to the prefrontal region. However, this motor system-based approach has been questioned over the years, as anatomical locations in the DLPFC have not been consistent (16, 17). Furthermore, the motor system may not be directly implicated in the disorder. In contrast, the default mode network (DMN), and the sgACC in particular, has been increasingly recognized as a region involved in both the pathophysiology (18, 19) and the clinical improvement of depression (20).

One approach to optimize coil placement utilizes resting state functional MRI connectivity of the individual. Building on evidence of a physiological anticorrelation between the frontoparietal network and the DMN (21), an rTMS study demonstrated that reducing the DMN hyperconnectivity in MDD, extending to the sgACC, was associated with symptomatic relief (18, 20). Subsequent studies have replicated an association between greater therapeutic success with rTMS and the degree of anticorrelation of the individual left DLPFC and a general sgACC seed (10). However, since a general sgACC seed may not reflect the individual sgACC site implicated in symptoms, we validated a protocol using the same anticorrelation concept, but with a focus on the strongest anticorrelation between the left dorsolateral prefrontal cortex (DLPFC) and the individual DMN. This approach demonstrated that rTMS resulted in functional disconnection of the DMN up to the sgACC region in healthy volunteers (22, 23). Translationally, we hypothesized that the closer the ideal stimulation site was to the individualized target, the greater the clinical benefit of the iTBS protocol would be for depressed patients and we tested both DMN and sgACC associations, as previously replicated.

A novel approach, termed neuro-cardiac-guided (NCG-)TMS, involves stimulating the left DLPFC and, possibly through the sgACC, exerting modulations on HR and HR variability. This is thought to reflect the successful activation of the frontal vagal pathway, which would enhance the vagal tone, reducing HR and/or increasing HR variability. Additionally, research in healthy volunteers has suggested that stimulation at the F3 position of the 10-20 EEG system is optimal for inducing early HR changes with rTMS (24). Compared to healthy controls, depressed patients exibit altered HR and HR variability (25, 26), and these alterations are not always reversed by treatment (27). However, another study using iTBS observed a trend toward an association between HR decelerations and clinical improvement in depression (8), suggesting that HR modulations may serve as a predictor for treatment response.

In the current RCT, we rigorously tested the two previously suggested approaches to enhance the clinical benefits of rTMS treatment in depression: (1) spatial customization of coil position in the left DLPFC based on resting-state functional connectivity data, and (2) early modulations of cardiac rhythm. The primary outcome was clinical improvement over a 6-week period, as assessed by the Montgomery-Åsberg Depression Rating Scale (MADRS) score in patients with major depression treated with accelerated iTBS. Based on the existing evidence, we hypothesized that both the personalized selection of stimulation sites (Personalized > F3 site) and the early iTBS-induced modulations of HR (as measured by RR slope) and HR variability change (as measured by RMSSD minus baseline) would be positively associated with greater clinical improvement.

## Methods

### Participants and Study Protocol

We selected 125 depressed patients between April 2019 and July 2021, recruited through national advertisements, to participate in an rTMS study. Inclusion criteria were age between 18 and 60 years, diagnosis of MDD confirmed on site by medical assessment using the Structured Clinical Interview for DSM-5 Disorders-Clinician Version (SCID-5-CV), and moderate to severe depressive episode, according to MADRS score. The intervention was on an outpatient basis, although both inpatient and outpatient participants were included. Exclusion criteria were epilepsy or other neurological diseases, pregnancy, metallic implants, claustrophobia, as contraindications to MRI or rTMS, or, in case of regular psychopharmacological medication intake, change of type or dosage in the 2 weeks prior to participation or during the course of the study. To account for the psychotropic medication load (28–32), we summed for each individual the estimated intake dose of each substance at each visit, based on the interview or when possible on the circulating blood levels (below = 1, normal =2, or above = 3 of the expected dose). The study protocol (clinicaltrials.gov/ct2/show/NCT05260086) is in accordance with the Declaration of Helsinki and was approved by the Ethics Committee of the University Medical Center Göttingen (UMG). Prior to enrollment, verbal and written informed consent was obtained from all participants.

Once enrolled, the participant was randomized to participate in a 6-week quadruple-blind (patient, stimulation provider, principle investigator, clinical rater) crossover trial. We used the randperm function in MATLAB (The MathWorks, Inc., Natick, MA, USA) to generate a true random sequence that coded if the participant would receive the stimulation either at the F3 position of the 10-20 EEG cap (F3 group) or at personalized left DLPFC sites (Personalized group) (Figure 1A), and either as active-sham or sham-active conditions (Figure 1B). At baseline and at the end of every week, a trained rater administered the MADRS (33) as a primary outcome measure, and additionally the Beck Depression Inventory (BDI-II) (34) and the Hamilton Depression Rating Scale (HAM-D) (35) as secondary outcome measures.

**Figure 1:**
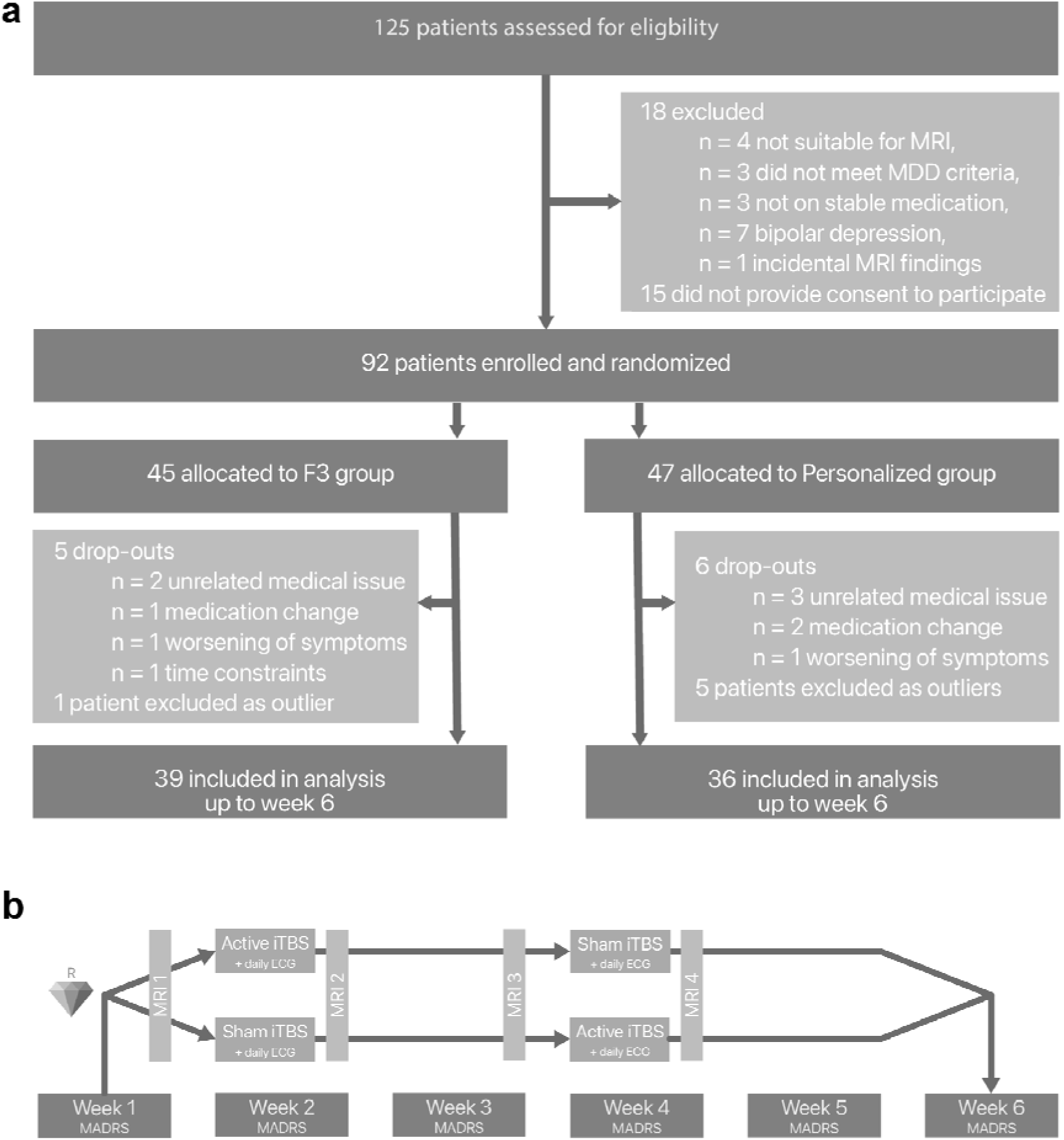
Participants and Study Protocol. (A) Study flow diagram and (B) study design

### Magnetic Resonance Imaging

We collected structural data with T1-weighted scans (1×1×1 mm) with a 64-channel head coil in a 3T MR scanner (MAGNETOM Prisma, Siemens Healthcare, Erlangen, Germany). We acquired the rsfMRI data using T2*-weighed gradient-echo echoplanar imaging with the following parameters (36, 37): repetition time of 1.5 s, echo time of 30 ms, flip angle of 70°, 69 axial slices with multiband factor of 3, 2×2×2 mm, FOV of 189 mm, with 10% interval between slices and posterior to anterior phase encoding. The rsfMRI data was acquired with 400 volumes in 10 minutes. The gradient-echo field map was acquired with a repetition time of 704 ms, echo times of 4.92 ms (TE 1) and 7.38 ms (TE 2), flip angle of 60°, 73 slices, FOV of 210 mm, 2×2×2 mm, with a 10% interval between cuts and phase encoding from anterior to posterior.

### Ideal and Stimulated Cortical Sites

In accordance with prior publications (22, 23), we temporally concatenated the data for group independent component analysis (GICA). The GICA was performed with FSL 5.0.7 software (38) separately for the scans of the first, second, third, and fourth week, and separately for the F3 and Personalized group. We identified components that best represented the DMN and the network that best covered the left DLPFC, and back reconstructed these into the normalized rsfMRI data of the individual patients. We then identified the left DLFPC region that most strongly anticorrelated to the DMN of the subject (ideal site) and projected the (x, y, z) coordinates from normalized to native space. Noteworthy, our method to determine the strongest left DLPFC anticorrelation to the DMN in each participant differs from a commonly used approach in which the area of strongest anticorrelation is found from seed-based connectivity analyses using a common sgACC seed derived from healthy controls (10). Therefore, we also extended the analysis to the seed method to explore whether this approach would yield different results.

Our neuronavigation systems (Visor 2 software, ANT Neuro, Enschede, Netherlands) recorded the stimulation coordinates in each session. We calculated the average of the first coordinate of each session in the week (x, y, z) and centered a region of interest (ROI) on it with spheres of 2 mm radius using MarsBar (39). To find the point on the cortex closest to the ROI on the skull, we calculated the Euclidean distance between the segmented cortex of each individual T1-weighted image and the center of the ROI using SPM12 and MATLAB. Finally, we calculated the Euclidean distance between the ideal cortical site and the stimulated cortical site for each observation and used this variable (“distance”) in further analyses.

### ECG Acquisition and Analysis

Participants were sitting quietly in a reclined chair for at least 10 min before recordings started. We acquired ECG data 2 min before (baseline) and throughout iTBS using the three electrodes of the neuroConn NCG-rTMS device (neuroCare, Munich, Germany) - ground electrode placed near the lower end of the sternum, red electrode placed on the 1st intercostal space on the right side, green electrode placed on the 4th or 5th intercostal space on the left side. The device recorded the ECG at 1000 Hz as well as the iTBS bursts, allowing for exact synchronization of ECG and iTBS information. We analyzed the ECG data using the Biosignal Processing in Python (BioSPPy) toolbox, the HRV analysis toolbox, and the SciPy module (40) in Python 3.6. We used SciPy to remove power-line noise from the ECG signal and then used BioSPPy to remove band wander noise in the data and detect R peaks of the QRS complex as the maxima of the curve ± 25 ms. Then, we calculated the RR interval using the timestamps of detected R-peaks. We then preprocessed the RR interval by developing a Python correction of ectopic beats and other types of outliers as detailed in Lipponen and Tarvainen, 2019 (41). Next, we selected the timeframe of 45 s from the start of iTBS for the RR interval, as the HR deceleration (represented by a positive slope) can be seen stronger in comparison to the sham condition (8). We also selected the root mean square of successive differences (RMSSD) in the time domain as an index of HRV due to being consistent and presenting the largest difference between MDD and HC among HRV parameters (42). The RMSSD is indicative of the parasympathetic influence of the autonomous nervous system on HR and lower values of RMSSD suggest lower parasympathetic (vagal) activity (43). Moreover, delta RMSSD showed significant decrease during cognitive stress (44). Based on criticism against ultra-short-term HRV and the recommendation of at least 4 min to draw accurate conclusions about HRV (45, 46), we selected the RMSSD timeframe of 270 s after stimulation starts.

### Accelerated Intermittent Theta Burst and Sham Stimulations

Stimulation sessions took place during week 2 and week 4. We used a MagVenture X100 with Mag-option and a figure-of-eight MCF-B65 A/P cooled coil. The resting motor threshold (RMT) was reassessed daily via electromyography in the right first dorsal interosseous muscle with the lowest intensity eliciting 50 mV motor reaction in 5 of 10 attempts, and the stimulation intensity was adjusted to 110%. In the active condition, we applied the stimulation parameters (4): stimulation ON lasted for 2 s, followed by 8 s of stimulation OFF, a volley composed of 10 units of three individual bursts with a frequency of 5 Hz and each burst with three single pulses with 50 Hz, performed 60 times, totalizing 1,800 per session (7,200 pulses daily and 36,000 pulses in total). In the sham condition, the same MCF-B65 A/P coil was blindly rotated 180 degrees, according to pre-coded sequences provided by the study PI, who had no contact with the participants. Each daily visit included four sessions (accelerated protocol) of approximately 10 minutes per session, followed by at least 20-minute pauses, according to the necessary inter-session time described on previous literature (47). As part of the device, tENS electrodes placed in the scalp as close as possible to the coil mimicked tingling skin sensations simultaneously to the sound of pulses, regardless of the stimulation condition. After completion of all stimulation sessions, participants were asked to indicate their expectation about the effectiveness of the stimulation using a visual analogue scale (VAS). As there was no significant difference between expectation levels under stimulation conditions (t(72) = 1.47, p = 0.15, see Table 1), we see the blinding procedure as effective.

**Table 1:** Characteristics of the participants. Demographic, clinical, and baseline characteristics of the participants included in the analysis.

|  | **Total** |  | **Personalized group** |  | **F3 group** |  |
| --- | --- | --- | --- | --- | --- | --- |
|  | N (%) | M ± SD | N (%) | M ± SD | N (%) | M ± SD |
| **Number of subjects** | 75 (100.00) |  | 36 (48.00) |  | 39 (52.00) |  |
| **Order active – sham** | 37 (49.33) |  | 17 (47.22) |  | 20 (51.28) |  |
| **sham – active** | 38 (50.67) |  | 19 (52.78) |  | 19 (48.72) |  |
| **Sex \*\*\*** |  |  |  |  |  |  |
| **male** | 42 (56.00) |  | 15 (41.67) |  | 27 (69.23) |  |
| **female** | 33 (44.00) |  | 21 (58.33) |  | 12 (30.77) |  |
| **Age** |  | 36.14 ± 13.09 |  | 33.83 ± 12.02 |  | 38.28 ± 13.74 |
| **Years of education** |  | 16.42 ± 3.48 |  | 16.70 ± 3.74 |  | 16.16 ± 3.24 |
| **Medication use** |  |  |  |  |  |  |
| **yes** | 57 (76.00) |  | 29 (80.56) |  | 28 (71.79) |  |
| **no** | 18 (24.00) |  | 7 (19.44) |  | 11 (28.21) |  |
| **Medication load** |  | 1.91 ± 1.77 |  | 2.13 ± 1.95 |  | 1.71 ± 1.58 |
| **MADRS at baseline** |  | 24.55 ± 7.26 |  | 24.32 ± 7.45 |  | 24.79 ± 7.09 |
| **Treatment resistance** |  |  |  |  |  |  |
| **yes** | 29 (38.67) |  | 18 (50.00) |  | 11 (28.21) |  |
| **no** | 46 (61.33) |  | 18 (50.00) |  | 28 (71.79) |  |
| **Number of subjects with comorbidities<sup>‡</sup>** | 20 (26.67) |  | 12 (33.33) |  | 8 (20.51) |  |
| **Presence of generalized anxiety** | 7 (9.33) |  | 3 (8.33) |  | 4 (10.26) |  |
| **panic disorder** | 3 (4.00) |  | - |  | 3 (7.69) |  |
| **eating disorder** | 2 (2.67) |  | 2 (5.56) |  | - |  |
| **social phobia** | 6 (8.00) |  | 3 (8.33) |  | 3 (7.69) |  |
| **obsessive compulsive disorder** | 2 (2.67) |  | 1 (2.78) |  | 1 (2.56) |  |
| **PTSD** | 2 (2.67) |  | 2 (5.56) |  | - |  |
| **somatiform** | 1 (1.33) |  | 1 (2.78) |  | - |  |
| **ADHD** | 1 (1.33) |  | 1 (2.78) |  | - |  |
| **Expectation of effect (VAS 0-1)** |  |  |  |  |  |  |
| **active** |  | 0.43 ± 0.29 |  | 0.45 ± 0.30 |  | 0.41 ± 0.28 |
| **sham** |  | 0.38 ± 0.29 |  | 0.39 ± 0.29 |  | 0.38 ± 0.30 |

<sup>‡</sup>Comorbidities ( $\chi^2(1) = 0.8, p = 0.371$ ): not included as covariates; \*\*\* $p < 0.001$

### Statistical Analysis

Statistical analyses were conducted in RStudio (48), adopting a significance level of two-sided α = 0.05. Participants were excluded from the analysis, if the baseline MADRS score < 7 or the z-standardized outcome score > ± 2.5 SD. Levene’s test was used to check the homogeneity of variance assumption and Mauchly’s test was used to check the sphericity of variances. To investigate sample characteristics in the F3 and Personalized groups, we used non-parametric Wilcoxon rank sum (W) and chi-square (χ2) tests. For investigating the MADRS scores and the interaction between groups over time, we conducted repeated mixed measures ANOVA (F) with the time as the within-subject factor and F3 or Personalized groups as a between-subjects factor. Due to significant differences between groups in sex distribution, we added sex as an additional between-subjects factor.

We conducted independent analyses of Wilcoxon rank sum (W) test between the F3 and Personalized groups to investigate the Euclidean distance. We further computed Pearson correlation coefficients between the distance and delta MADRS as well as between HR/HRV and delta MADRS. By conducting dependent t-test (t) or Wilcoxon rank sum (W) test, if the assumption of normality in the data is violated, we compared the ECG measures between the active and the sham condition. We calculated Cohen’s *d* and the 95% confidence interval as effect size within and between groups and conditions. An effect size of *d* > 0.10 indicates a small, an effect size of *d* > 0.30 a medium, and an effect size of *d* > 0.50 a large effect (49). Delta MADRS score was computed as *1 – post-stimulation score / pre-stimulation score*. The primary outcome was delta MADRS score within 6 weeks, but we also investigated delta MADRS score within each stimulation week as an attempt to unravel possible carryover effects. For the later, we used the R packages lme4 (50) and lmerTest (51) to fit the linear mixed-effects models (LMM) using the restricted maximum likelihood (REML) method and to extract p values. We performed full-null comparisons using the chi-square (χ2) test of our hypothesized full model and a corresponding null model with a random structure. To minimize the accumulation of α error, we only extracted and interpreted the estimated fixed effects when the full model was significantly different from the null model.

## Results

Of the 125 patients screened, 92 were enrolled in the study (Figure 1). Of the 33 patients who were not enrolled, 18 were excluded and 15 did not consent to participate, mainly due to time constraints. Of the excluded patients, 7 met the diagnostic criteria for bipolar depression, 3 did not meet the diagnostic criteria for MDD, 4 had claustrophobia, 3 were on recently adjusted medication, and 1 presented an incidental finding in the MRI scan. Of the 92 participants, 11 dropped-out during the 6-week course of the study, with no reports of serious adverse effects (see Safety section below). The reasons for dropping-out of the study were rTMS-unrelated issues (e.g., symptoms of a cold), time limitations, change in medication, etc. We also excluded 6 patients from the final analysis due to errors in the ECG acquisition (incomplete acquisition, or extreme values detected as outliers), resulting in 75 participants.

### Characteristics of the Sample

We assessed the age, sex, years of education, medication use, treatment resistance (TRD), comorbidities, and expectation of effect (blinding) of the participants in total and in each group (Table 1). Except for sex (*χ*2(1) = 10.45, *p* = 0.001), no other characteristic was different between groups. The number of patients on stable medication did not differ between groups (*χ*2(1) = 0.03, *p* = 0.872).

### Safety and Side Effects

Of the 75 participants, 53.33% reported headache, 30.00% neck pain, 58.00% scalp pain, and 20.67% scalp irritation at the stimulation site. The intensity of daily pain/irritation experienced (Likert 0-10) was, however, very low on average in the active condition (M(SD): 1.47 (1.19) headache, 1.31 (1.35) neck pain, 2.31 (1.73) scalp pain, 1.40 (1.03) scalp irritation) as well as in the sham condition (M(SD): 1.01 (0.99) headache, 1.24 (1.32) neck pain, 1.06 (0.90) scalp pain, 0.81 (0.55) scalp irritation). The intensity of daily pain/irritation between conditions differed (*p < 0.0125*) only for headache (W = 3663.5 p= 0.007 for headache, W = 2688 p = 0.564 neck pain, W = 3369.5 p = 0.029 scalp pain, W = 2744.5 p = 0.718 scalp irritation). No participant reported serious adverse effects (SAE).

### Personalized vs F3 Stimulation

We found no significant interaction effect of time and sex in the two groups (F(4,295) = 0.733, p = 0.57). Furthermore, no two-way interactions were significant. There was a significant effect of time (F(4,295) = 14.378, p < 0.001). In the pairwise comparisons, a significant reduction of MADRS was found in F3 from V1 to all visits except V3, whereas in the Personalized, only V6 showed a significant reduction compared to V1 after Bonferroni correction. Most importantly, patients in the F3 when compared to the Personalized stimulation displayed no significant difference in MADRS scores at the first as well as the last visit (pairwise comparison p = 0.494, p = 0.490) (Figure 2 Panel 1). Therefore, there was a significant reduction in MADRS scores in both groups, but no significant difference between personalized and standard F3 stimulation was found in the V6 evaluation. Finally, although difficult to visualize, the within-week reductions in the overall MADRS score in the active condition were significantly larger than the sham in the LMM (Table 2), as seen earlier in (45).

**Figure 2:**
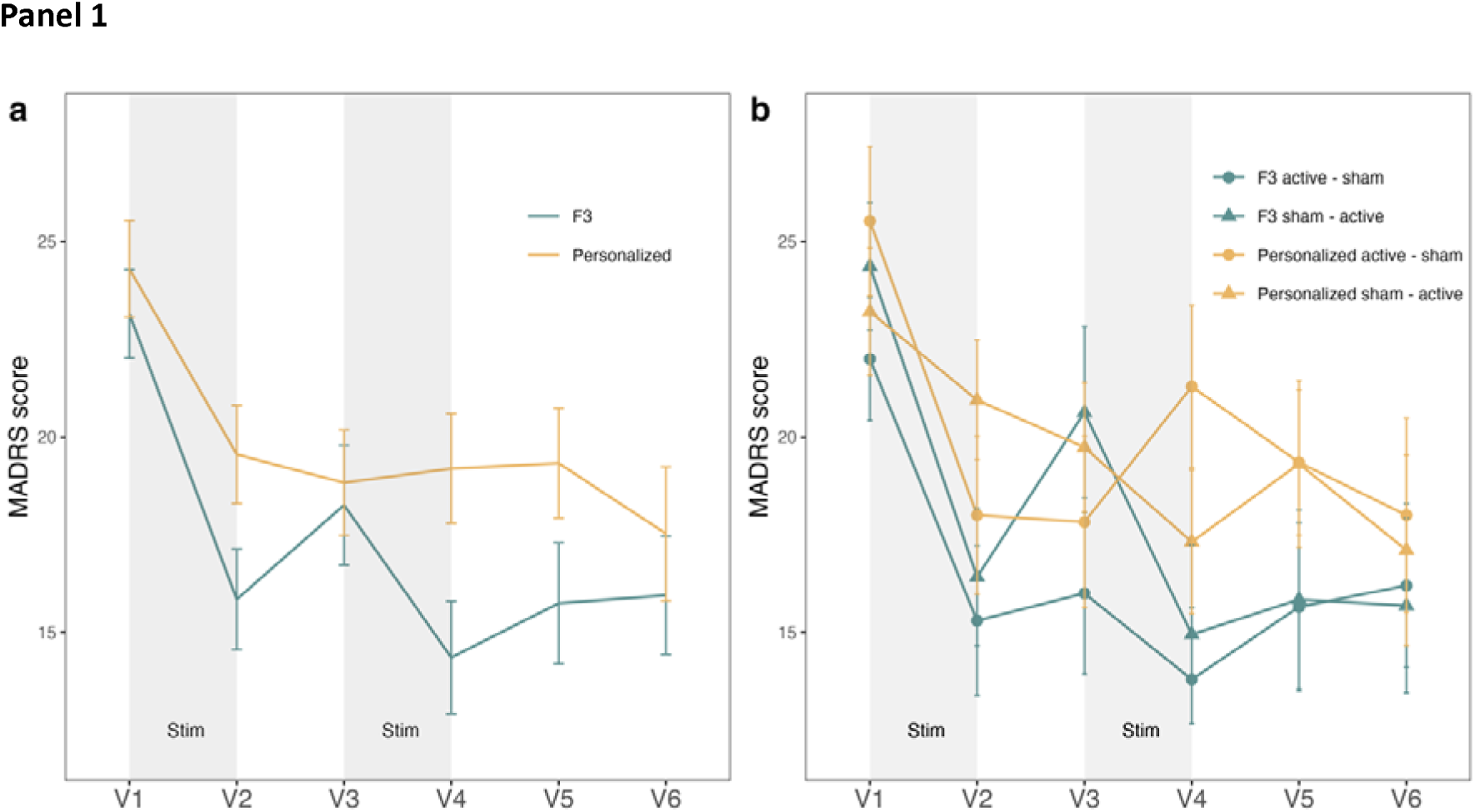
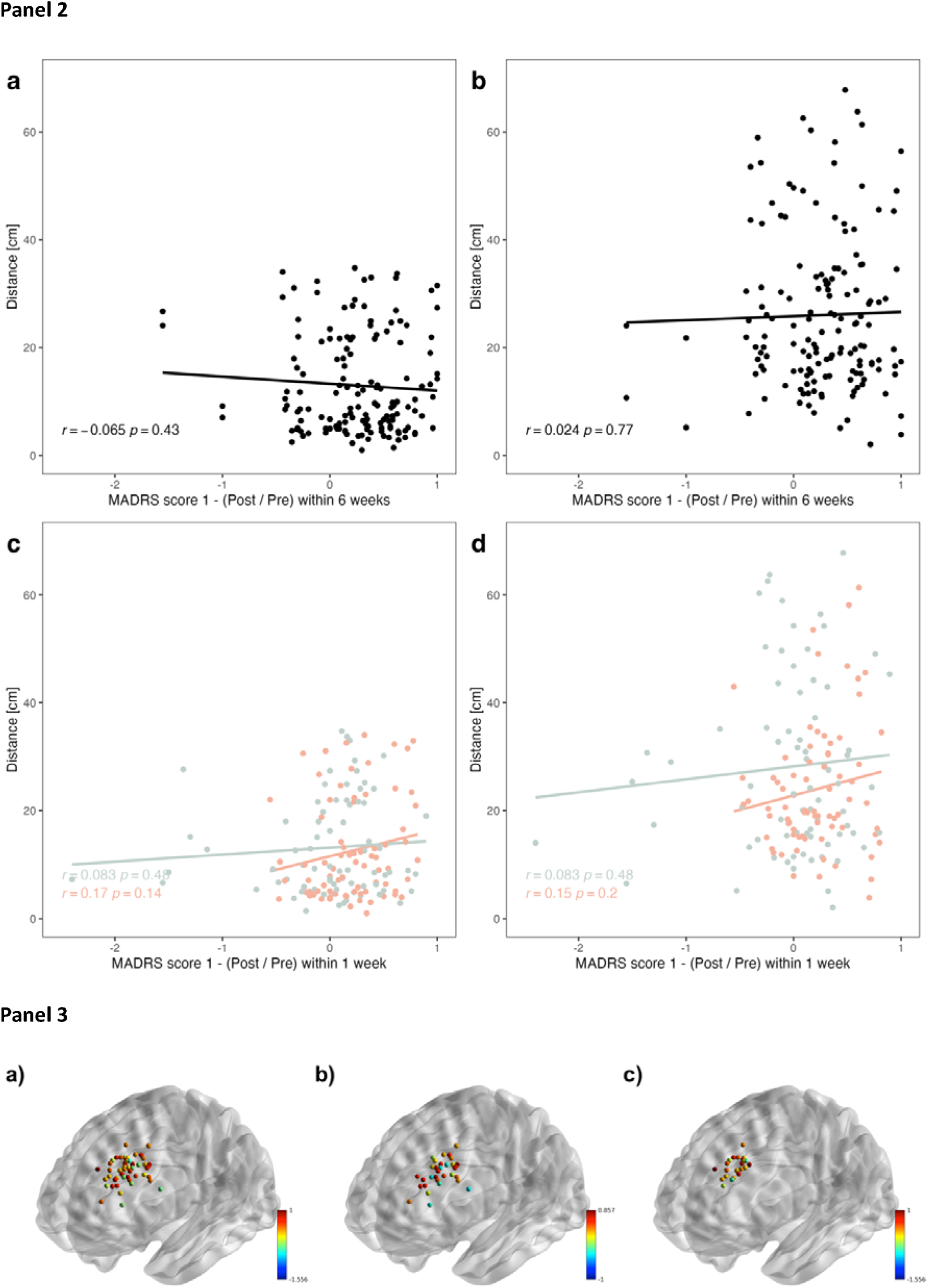
MADRS Scores, Stimulation Site Distance, and Spatial Distribution in F3 and Personalized Groups. Panel 1) Montgomery–Åsberg Depression Scale (MADRS) score from weekly visits V1 to V6 with active or sham stimulation between V1-V2 and V3-V4 visits (A) in F3 and Personalized groups and (B) in F3 and Personalized groups with sham-active and active-sham subgroups. Error bars represent mean ± SE. Panel 2) The relationship between the clinical improvement based on delta MADRS score and the distance to ideal stimulation site calculated by (A,C) the default mode network (22, 23) and (B,D) the subgenual anterior cingulate cortex (9–11) methods. Note active (red) vs sham (gray) in (C,D) Panel 3) Individual stimulation sites in native space projected to the MNI space in (A) whole sample (B) personalized and (C) F3 groups. Points are color-coded according to MADRS score 1 – (Post / Pre) within 6 weeks. Note the absence of a clear spatial pattern of color distribution.

**Table 2:**
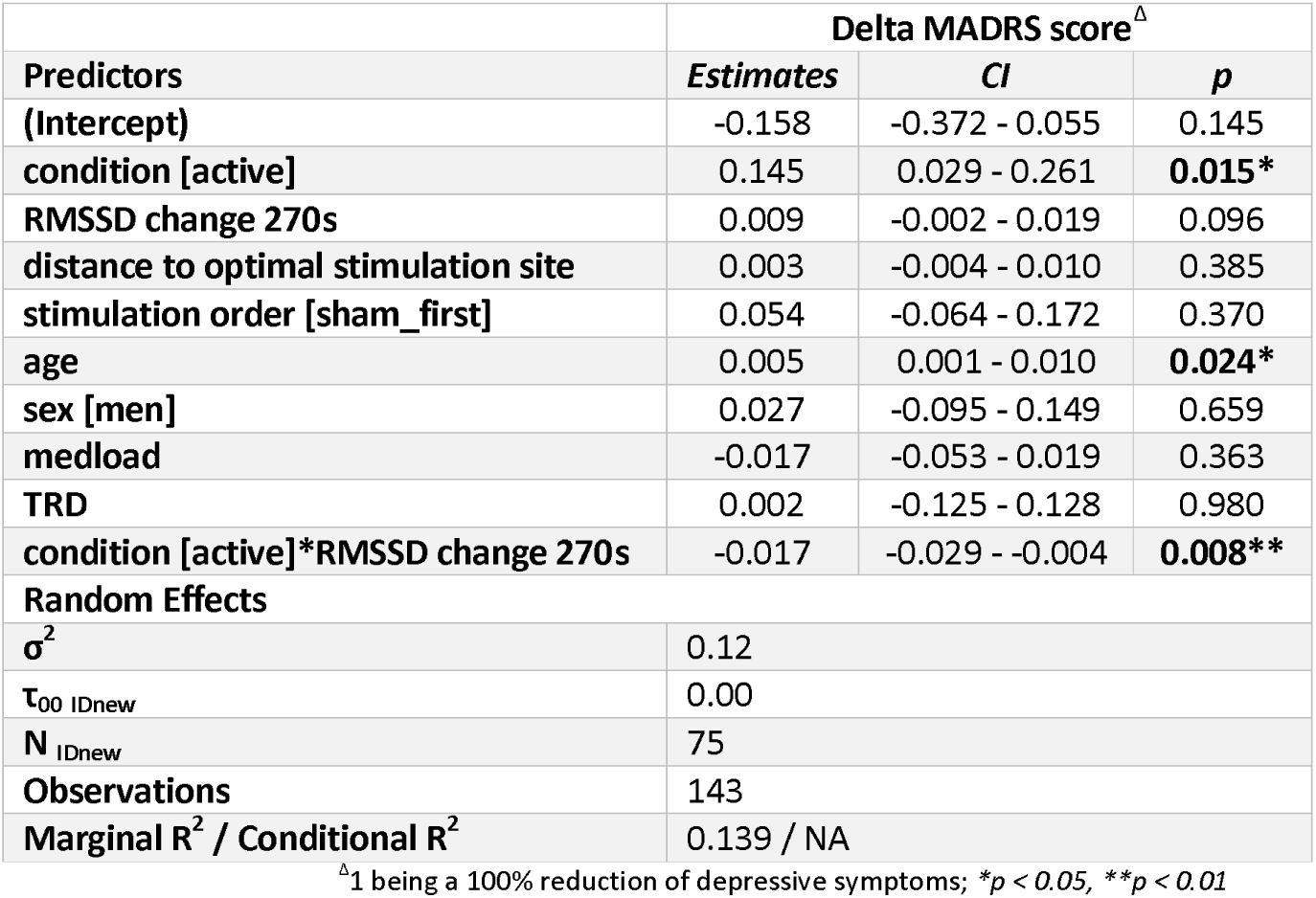
Linear mixed model analysis of the predictors of changes in Montgomery–Åsberg Depression Scale (MADRS) scores.

Next, we investigated whether the MADRS score changed according to the distance between the stimulated and ideal sites. The mean Euclidean distance between ideal and stimulated sites was obviously shorter (W = 4826, p < 0.001) in the Personalized (M ± SD: 7.27 ± 4.76 mm) as compared to the F3 (M ± SD: 18.20 ± 9.17 mm) group. However, the correlation values for active (r = 0.17, p = 0.14) and sham (r =0.08, p = 0.48) conditions did not hint towards any relationship (Figure 2 Panel 2). In addition, we projected the stimulation sites from the native to the standard space in order to visualize the spatial distribution of the areas in Personalized and F3 groups coded with clinical response ranges (Figure 2 Panel 3).

### Heart Rate and Heart Rate Variability

We assessed whether iTBS-driven modulations in the RR interval (45 s) and RMSSD (270 s) would be associated with clinical benefit (delta MADRS score: 1 being a 100% reduction of depressive symptoms). Therefore, we correlated slopes and delta RMSSD with MADRS change and found a significant positive relationship for slope in the active condition only (r = 0.27, p = 0.021, Figure 3A), which was also significantly higher than the sham condition (t(74) = 2.92, p = 0.005, d = 0.41 (CI: 0.12 – 0.69), Figure 3B). A relationship was not seen for correlation of delta MADRS score and RMSSD change in the active (r = -0.17, p = 0.15) or sham (r = 0.05, p = 0.7) conditions. When assessing the delta MADRS within a week, we identified a significant negative correlation with RMSSD change in the active condition only (r = -0.29, p = 0.013, Figure 3C), which was also significantly higher than the sham condition (W = 3402, p = 0.017) (Figure 3D). To confirm that the hypothesized timeframes were correct, we also conducted an exploratory investigation of timeframes 30 s, 45 s, 60 s, 180 s, 270 s, and 360 s to cover time regularly up to 6 min (Tables S1, S2, and S3, Figures S1 and S2).

**Figure 3:**
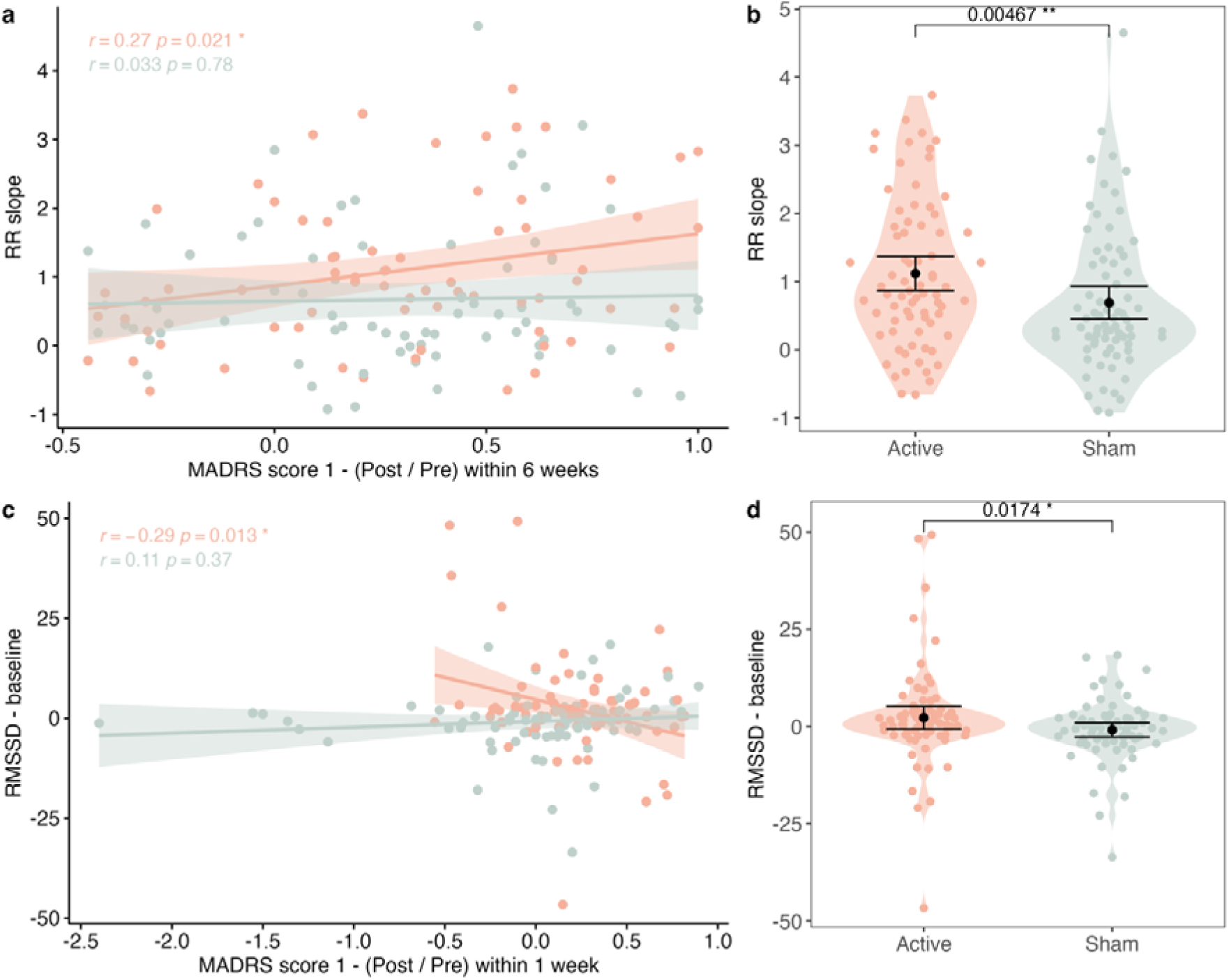
Stimulation-induced the RR interval (first 45 seconds) and RMSSD (first 270 seconds) modulations along with the corresponding Montgomery–Åsberg Depression Scale (MADRS) score changes. (A) The association between the RR slope and delta MADRS score within 6 weeks was seen in the active condition (red) only, and (B) the RR slope was stronger in active (red) vs sham (gray) conditions. (C) The association between the RMMSD change and delta MADRS score within 1 week was seen in the active condition (red) only, and (D) the RMMSD change was stronger in active (red) vs sham (gray) within 1 week. Error bars represent mean ± SE. *p < 0.05, **p < 0.01

Additionally, we performed LMM to explore the within-week effect of iTBS on delta MADRS score, which, in the context of hypothesis-generating, was not controlled for multiple testing. The first set of models with the three-way interaction between the condition, the distance, and the RR slope yielded a significant full-null comparison at 45 s (χ2(6) = 13.09, p = 0.042). At all other timeframes (30 s, 60 s, 180 s, 270 s, 360 s), the full model was not significantly different from the null model. We consequently calculated the models for 45 s only. The three-way interaction was not significant (p > 0.05). We decided to reduce the model to the two-way interactions between the condition and the distance, the condition and the slope, and the distance and the slope. Again, we did not observe any significant interaction. We further reduced the 45 s model to the main effects and observed a significant effect for the age predictor (β = 0.006 p = 0.012). As the full-null comparisons did not yield any significant differences (p > 0.05), we could not replicate this finding with our secondary outcome measures (HAMD, BDI-II) as the dependent variables.

The second set of models with the three-way interaction between the condition, the distance, and the RMSSD difference score was significantly different from the null model at all timeframes [χ2(6) = 12.70, p = 0.048 (30 s), χ2(6) = 14.83, p = 0.021 (45 s), χ2(6) = 14.19, p = 0.028 (60 s), χ2(6) = 13.81, p = 0.032 (180 s), χ2(6) = 17.58, p = 0.007 (270 s), and χ2(6) = 24.67, p = 0.0004 (360 s)]. Still, the three-way interaction was not significant (p > 0.05). We reduced the models by removing the insignificant interactions between the condition and the distance, and between the RMSSD change and the distance. In doing so, the two-way interaction was significant in the models for 180 s (β = -0.015, p = 0.011) and for 270 s (β = -0.017, p = 0.008, Table 2). We saw a trend in the model for 60 s (β = -0.01, p = 0.075). These estimated fixed effects were all negative, indicating that a greater iTBS-specific change in RMSSD led to worse treatment outcome in our sample. Regarding our secondary outcomes, none of the full null comparisons were significant in HAMD. The set of models with BDI-II was significantly different from the null model at 180s (χ2(6) = 22.31, p = 0.001), 270s (χ2(6) = 19.76, p = 0.003) and 360s (χ2(6) = 21.19, p = 0.002). The three-way interactions were all insignificant. However, the two-way interactions between RMSSD change and active condition were significant for 180s (β = -0.022, p < 0.0001), 270s (β = -0.020, p = 0.002) and 360s (β = -0.022, p = 0.001).

## Discussion

In this RCT, we investigated whether the clinical benefit of accelerated iTBS for treating depression could be related to personalization of left DLPFC stimulation sites and/or early heart rhythm modulations. As the largest prospective study to date to address these aspects, our findings show that clinical improvement driven by iTBS was associated with modulations of HR and HR variability - but not with personalized stimulation sites based on individual resting state connectivity. As hypothesized, iTBS-driven modulations of HR were significantly associated with clinical improvement in depression. Specifically, greater HR deceleration (RR slope) within the first 45 s of the first stimulation day, the greater the clinical benefit, reflected in MADRS score changes up to 6 weeks of follow-up. Although iTBS-induced HR variability changes (RMSSD) within the first 270 seconds did not correlate with MADRS score change at 6 weeks, they did predict clinical benefit at a shorter follow-up (1 week), with patients showing lower RMSSD change experiencing greater MADRS score reductions. However, no clinical benefit was observed from using personalized stimulation sites based on individual functional connectivity (Personalized site) compared to fixed stimulation sites (F3 site), nor was a relationship found between the distance from ideal sites on the left DLPFC and clinical improvement.

There is considerable anticipation regarding the identification of methods that maximize the therapeutic effect of rTMS (52, 53). The neuro-cardiac-guided-TMS approach suggests that when the frontal vagal pathways is subjected to high-frequency stimulation, HR deceleration can be observed, as demonstrated in healthy volunteers (24). In the context of MDD, often characterized by elevated HR (54), the HR deceleration observed early in the stimulation protocol could serve as objective evidence for potential effectiveness of the treatment. If this modulation in certain individuals is reactive, it possibly indicates successful activation of the frontal vagal pathway, thereby providing a means to predict which patients will benefit from the intervention. Our results support observation of increased RR slopes under real versus sham iTBS conditions, along with significant correlations between RR slopes and treatment improvement—findings that were previously only observed as trends (8). In fact, this approach has been regarded as so promising that some researchers have suggested it could replace the use of motor output to determine individual stimulation intensity and/or coil positioning in MDD patients (55). While the use of motor output to determine individual stimulation intensity is a well-established and valuable method for standardization, a clear link between motor cortex action potential thresholds and the left DLPFC remains uncertain. This raises concerns about the accuracy of transferring motor output intensity information to the level required for therapeutic effects. In other words, a model that considers the individual anatomy and function of the DLPFC would be more accurate, as the intensity determined in the motor cortex is dependent on its spatial characteristics, which may differ significantly from those of therapeutic targets. Therefore, a method grounded in the underlying mechanism by which appropriate stimulation of the left DLPFC probably targets the sgACC and triggers cardiac modulations appears highly promising for future clinical applications.

In addition, a meta-analysis identified lower RMSSD as a strong pathological finding in MDD in comparison to HC (42). Generally, HR variability is considered an indicator of well-being (56) and self-regulatory ability (12, 57), both of which are reduced in individuals with depression (42, 58). In this sense, an increase in RMSSD during iTBS could indicate greater frontal vagal modulations, from the DLPFC, through the sgACC to the brainstem and vagus nerve, reaching the heart (52), and thus serving as a potential predictor of clinical improvement. However, to the best of our knowledge, this mechanism has not been demonstrated in previous studies. While we were able to replicate the increase in RMSSD in active versus sham iTBS (8, 59), our exploratory results revealed an unexpected negative relationship. We found that the smaller the change in RMSSD, the greater the reduction in MADRS score. These findings, derived from a linear mixed model (LMM), show a significant two-way interaction between the iTBS condition and RMSSD change in predicting delta MADRS score. To further investigate these early heart rhythm modulations, we conducted additional analyses. A direct comparison of baseline MADRS scores between two subgroups, divided by the median RMSSD, showed no significant differences (p > 0.05). This suggests that our results were not influenced by potential flooring effects (e.g., less depressed individuals with higher RMSSD having less room for improvement compared to more depressed subjects). Based on the negative association, we hypothesized that effective engagement of the frontal vagal network would initially result in less HR variability during stimulation, followed by increased HR variability afterwards as a compensatory mechanism, aligning with clinical improvements. We further examined whether participants who improved clinically were also those who exhibited higher RR slopes and lower RMSSD changes at different time points. Of the 10 participants with the most significant changes, only 2 showed the greatest clinical improvement, indicating that more complex mechanisms may underlie these modulations. Overall, our results underscore the complexity of RMSSD as an output (60), suggesting that systematic investigation is needed to better delineate the brain-heart pathways and potentially achieve even higher predictive accuracy (8) than that observed with RR slopes alone.

Equally important, regarding clinical improvement, inter-individual variability could potentially be reduced, and the effect of iTBS maximized, by optimizing stimulation sites based on individual functional connectivity data. The sgACC has been proposed as a key marker for treatment response in depression (9, 20), and several studies have suggested that stimulation sites with the strongest negative connectivity to the sgACC could yield superior clinical outcomes (10, 13, 14). This method has been widely replicated and is considered the most established approach in the current literature for identifying patients who may benefit from rTMS. However, in our study, the use of functional connectivity data for personalized site selection did not show a clinical advantage. When comparing personalized stimulation sites on the left DLPFC to the fixed F3 stimulation site, no significant difference in MADRS score changes at 6 weeks of follow-up was observed. Additionally, no association between the distance from the ideal stimulation site and clinical improvement was identified. An inspection of the data revealed no spatial pattern of stimulation in the left DLPFC that suggested a clinical advantage. Importantly, because the point on the left DLPFC with the strongest anticorrelation to each subject’s DMN differs from the point with the strongest anticorrelation to a general sgACC seed, we repeated the analysis as described in previous studies (10). Again, no relationship between the distance from the ideal site and clinical improvement at 6 weeks was found (Figure 2) with accelerated iTBS. This finding contradicts the results of prior studies and may suggest that the profile of the MDD sample evaluated plays a critical role. Given that our study included a larger sample than those in previous studies showing positive results (9, 10), a sample size limitation is unlikely to explain the discrepancy. We also explored the changes in stimulation conditions within one week of the intervention but again found no evidence of a relationship. It is possible, however, that subgroups, such as biotypes (61, 62), may respond differently to this personalized approach, leading to better outcomes. Nevertheless, as it stands, we could not demonstrate any clinical benefit from the implementation of this personalization method. Our results align with Morriss et al. (63), with the advantage that those authors compared rsfMRI-with MRI-guided protocols and found no difference in clinical benefit, whereas we compared it with the F3 position of the EEG 10/20 system — an approach that is much simpler and more accessible in clinical practice. Overall, it must be acknowledged that using resting state functional connectivity data as the basis for this precision method was labor-intensive, expensive, and did not reveal a direct clinical advantage in this RCT. Future novel approaches (64) may provide new insights into this challenge.

Regarding the secondary outcomes (BDI-II and HAMD), a previous study found a negative correlation between baseline RMSSD and baseline HAMD-24 (65), which was not observed in our sample using the HAMD-17. Other findings could not be replicated, except for the two-way interactions between RMSSD change and the active condition in delta BDI-II score LMM models.

The present study has several limitations. The heterogeneity of depression, due to variations in presenting symptoms, is well-documented, prompting efforts to identify subtypes and their underlying neural correlates (61, 66, 62, 67). Due to limitations in sample size and insufficient representation of MDD clinical variations, we were unable to explore these important aspects. This should be a focus for future studies with larger and more diverse datasets. Additionally, we found no relationship between RR slopes or RMSSD changes and delta BDI-II or HAMD scores. This may be explained by the factor analytic approach of Uher et al. (68), who showed that the MADRS, BDI-II, and HAMD items load differently on three factors: the MADRS mainly on the Observed Mood factor, the BDI-II on the Cognitive factor, and the HAMD on the Neurovegetative factor. Given the dimensional complexity of MDD, it is not surprising that clinical improvement is not captured equally across all depression scales. Another limitation is the crossover design, which complicates the ability to separate effects between the iTBS conditions in the long-term assessment due to potential carryover effects from the active to the sham condition. This issue has been extensively discussed in another publication (69). Furthermore, although placebo effects were not sustained, they were robust, indicating that the rTMS setup may inflate expectation effects, as debated in the literature (70, 71) Thus, it is crucial for future studies to control for such effects as best as possible.

In conclusion, we demonstrated an association between HR deceleration during the first 45 seconds of stimulation and the degree of reduction in depressive symptoms up to 6 weeks after the aiTBS protocol. However, this relationship was not observed in HR variability modulation at 270 seconds, which correlated with clinical improvement only within the stimulation week, presenting important predictive limitations. Moreover, clinical improvement was not related to the distance between stimulation and ideal sites, and no benefit was observed between the Personalized and F3 groups. In contrast, the use of online ECG emerges as a promising and cost-effective approach that could be further developed for MDD treatment with iTBS. Overall, the promise of precision medicine may be realized through improved targeted stimulation using a combination of neuronavigation and ECG, which could allow for the stratification of patients more likely to benefit from iTBS.

## Data availability

The data that support the findings of this study are available from the corresponding author upon reasonable request.

## Acknowledgements

We thank the study participants and the colleagues from the Laboratory of Systems Neuroscience and Imaging in Psychiatry, the Department of Psychiatry and Psychotherapy, and the Department of Cognitive Neurology of the University Medical Center Göttingen. In particular, we are grateful for the great dedication of Ms. Ilona Pfahlert and Ms. Britta Perl. We thank Dr. Roger Mundry from the Leibniz ScienceCampus Primate Cognition (https://www.primate-cognition.eu/en) for statistical support. We also thank Mr. Klaus Schellhorn, Dr. Farnoosh Safavi and Mr. Matthias Kienle on behalf of customer service at neuroConn, ANTNeuro and MagVenture, respectively. It is important to note that customer service had no influence on the work described here. This work was supported by the German Federal Ministry of Education and Research (Bundesministerium für Bildung und Forschung, BMBF: 01 ZX 1507, ‘‘PreNeSt - e:Med’’). Jonas Wilkening was supported by the Göttingen Promotionskolleg für Medizinstudierende, funded by the Jacob-Henle-Programm/Else-Kröner-Fresenius-Stiftung and Jens Wiltfang is supported by an Ilídio Pinho Professorship, iBiMED (UID/BIM/04501/2013) and FCT project PTDC/DTP-PIC/5587/2014 at the University of Aveiro, Portugal.

## Supplementary Material for

**Table S1:** The mean RR slope values for the entire sample as well as separately for the active and the sham stimulations at baseline, 30, 45, 60, 180, 270, and 360 s timeframes. Standard deviations are specified in parentheses. The difference between conditions (active minus sham) is listed in the last column with *p* values of the dependent t test.

| Timeframe | Total | Active | Sham | Difference between conditions |
| --- | --- | --- | --- | --- |
| Baseline | -0.04 ± 0.39 | -0.13 ± 0.39 | 0.05 ± 0.39 | -0.08 (*t*(74) = -2.70, *p* = 0.008**) |
| 30 s | 1.11 ± 1.29 | 1.28 ± 1.32 | 0.92 ± 1.24 | 0.36 (*t*(74) = -2.46, *p* = 0.016*) |
| 45 s | 0.93 ± 1.10 | 1.14 ± 1.11 | 0.72 (1.06) | 0.42 (*t*(74) = -2.92, *p* = 0.005**) |
| 60 s | 0.65 ± 0.88 | 0.89 ± 0.95 | 0.42 ± 0.81 | 0.47 (*t*(74) = -2.52, *p* = 0.002**) |
| 180 s | 0.05 ± 0.23 | 0.09 ± 0.24 | 0.01 ± 0.22 | 0.08 (*t*(74) = -2.06, *p* = 0.043*) |
| 270 s | 0.01 ± 0.15 | 0.03 ± 0.16 | -0.02 ± 0.08 | 0.05 (*t*(74) = -1.76, *p* = 0.082) |
| 360 s | -0.01 ± 0.11 | 0.00 ± 0.13 | -0.02 ± 0.08 | 0.02 (*t*(74) = -1.44, *p* = 0.154) |

\**p* < 0.05

**Table S2:**
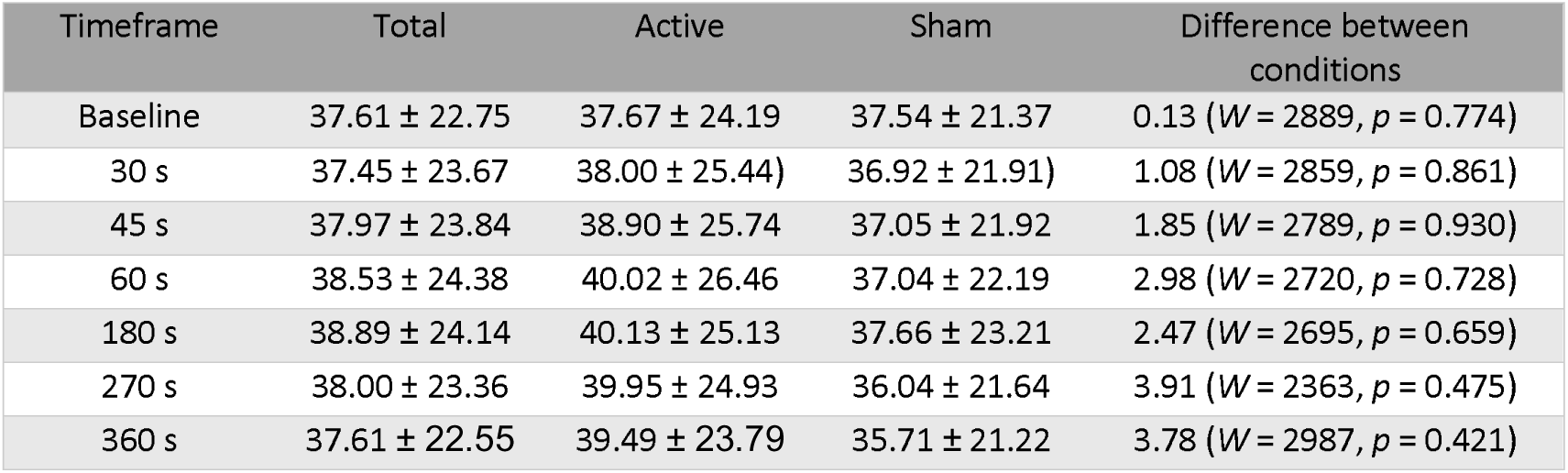
The mean RMSSD values for the entire sample as well as separately for the active and the sham stimulations at baseline, 30, 45, 60, 180, 270, and 360 s timeframes. Standard deviations are specified in parentheses. The difference between conditions (active minus sham) is listed in the last column with *p* values of the Wilcoxon test.

**Table S3:** The RMSSD change (mean RMSSD-baseline) scores for the entire sample as well as separately for the active and the sham stimulations at 30, 45, and 60, 180, 270, and 360 s timeframes. Standard deviations are specified in parentheses. The difference between conditions (active minus sham) is listed in the last column with *p*-values of the Wilcoxon test.

| Timeframe | Total | Active | Sham | Difference between conditions |
| --- | --- | --- | --- | --- |
| 30 s | -0.15 ± 11.45 | 0.33 ± 11.24 | -0.62 ± 11.72 | 0.48 (*W* = 2961, *p* = 0.577) |
| 45 s | 0.37 ± 10.83 | 1.22 ± 10.20 | -0.49 ± 11.43 | 1.57 (*W* = 3069, *p* = 0.335) |
| 60 s | 0.92 ± 11.65 | 2.35 ± 11.59 | -0.49 ± 11.61 | 2.47 (*W* = 3211, *p* = 0.134) |
| 180 s | 1.29 ± 10.77 | 2.46 ± 12.70 | 0.11 ± 8.34 | 2.80 (*W* = 3555, *p* = 0.041*) |
| 270 s | 0.72 ± 10.62 | 2.28 ± 12.71 | -0.87 ± 7.74 | 3.43 (*W* = 3402, *p* = 0.017*) |
| 360 s | 0.32 ± 10.56 | 1.81 ± 12.62 | -1.20 ± 7.74 | 3.21 (*W* = 3371, *p* = 0.024*) |

\**p* < 0.05

**Figure S1:**
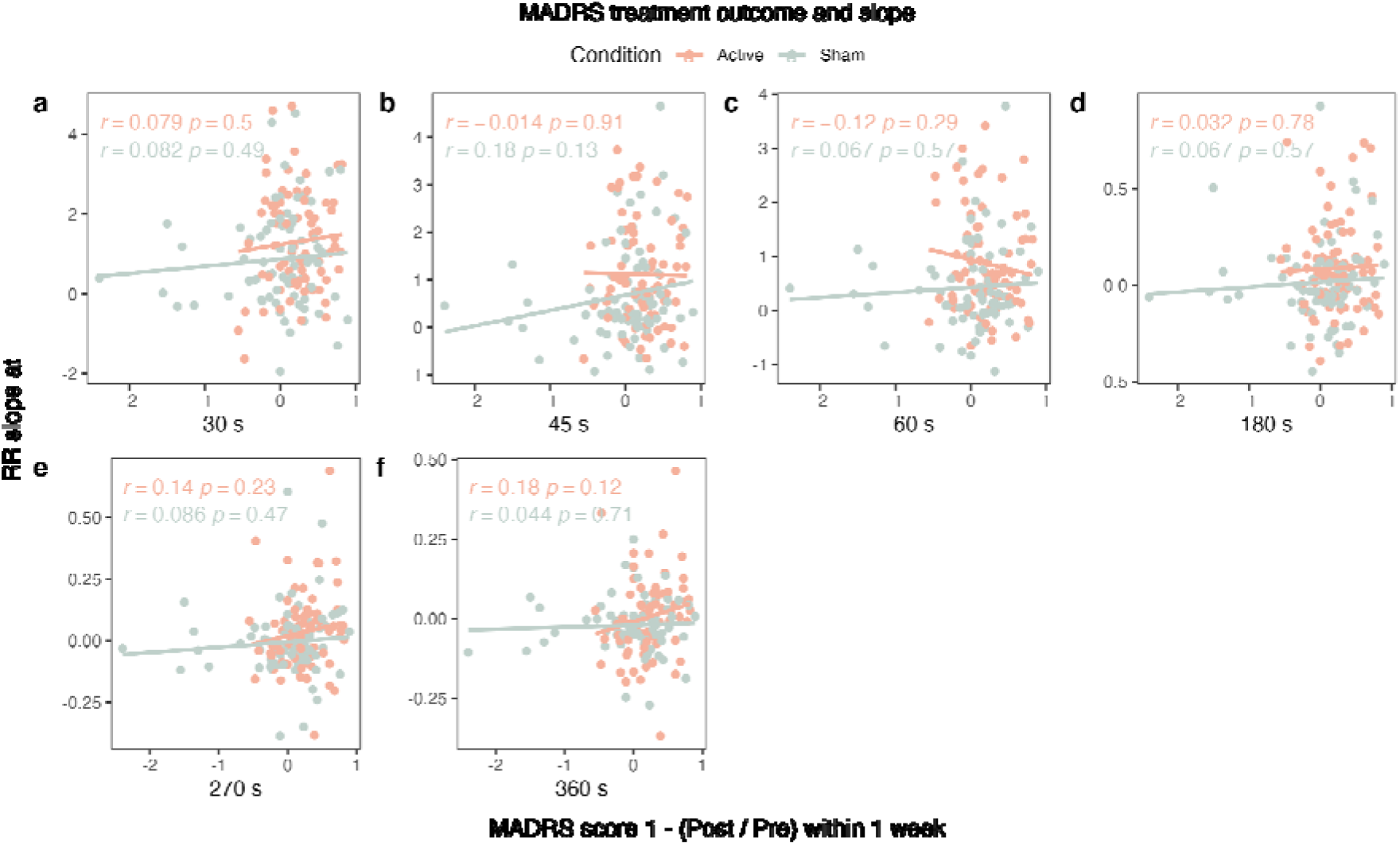
The relationship between the RR slope and delta MADRS score within one week was explored under active (red) and sham (gray) conditions over timeframes of (A) 30 s, (B) 45 s, (C) 60 s, (D) 180 s, (E) 270 s, and (F) 360 s from stimulation start.

**Figure S2:**
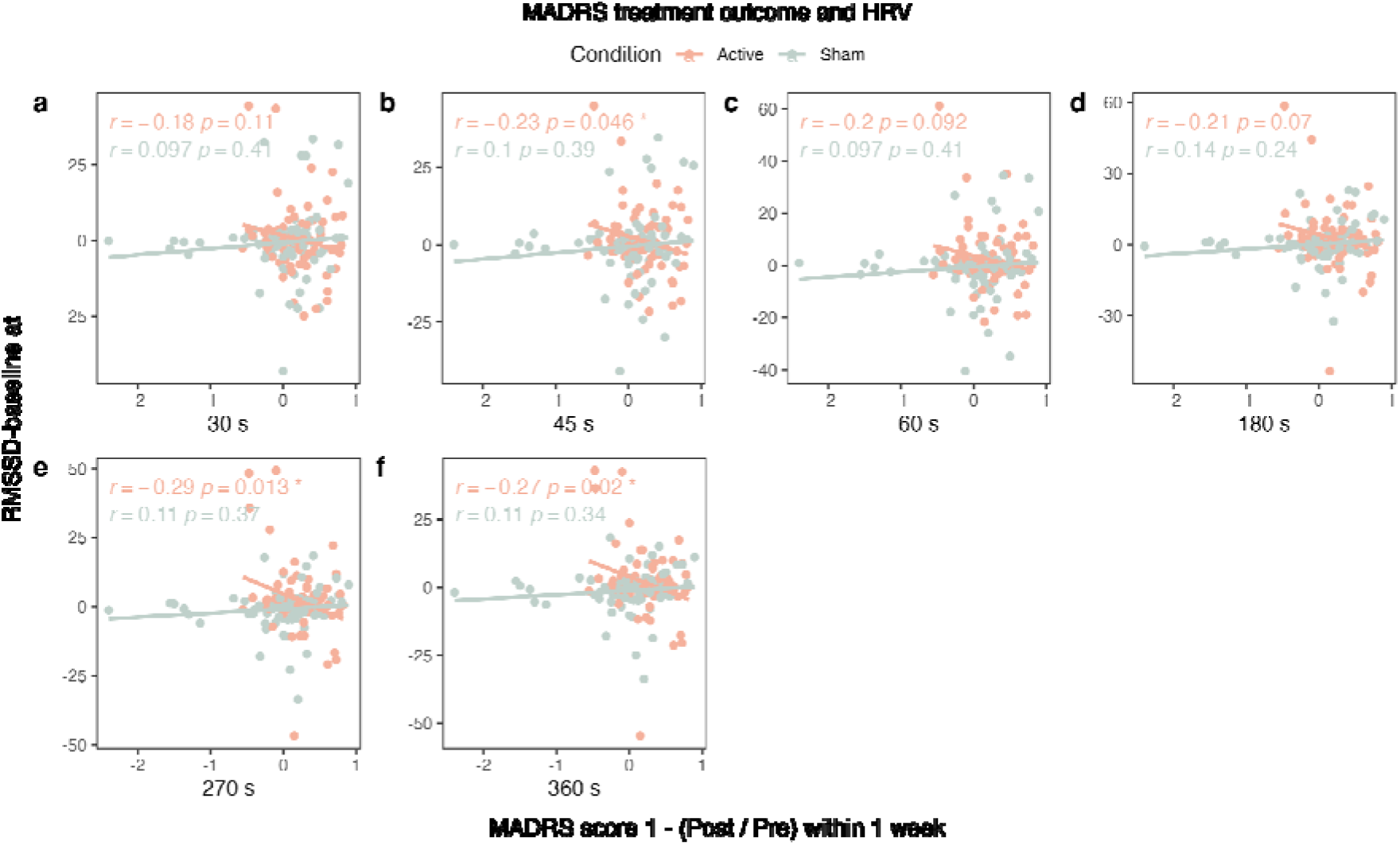
The relationship between the RMSSD change (RMSSD - baseline) and delta MADRS score within one week was explored under active (red) and sham (gray) conditions over timeframes of (A) 30 s, (B) 45 s, (C) 60 s, (D) 180 s, (E) 270 s, and (F) 360 s from stimulation start.

